# Machine Learning Approach for Confirmation of COVID-19 Cases: Positive, Negative, Death and Release

**DOI:** 10.1101/2020.03.25.20043505

**Authors:** Shawni Dutta, Samir Kumar Bandyopadhyay

## Abstract

In recent days, Covid-19 coronavirus has been an immense impact on social, economic fields in the world. The objective of this study determines if it is feasible to use machine learning method to evaluate how much prediction results are close to original data related to Confirmed-Negative-Released-Death cases of Covid-19. For this purpose, a verification method is proposed in this paper that uses the concept of Deep-learning Neural Network. In this framework, Long short-term memory (LSTM) and Gated Recurrent Unit (GRU) are also assimilated finally for training the dataset and the prediction results are tally with the results predicted by clinical doctors. The prediction results are validated against the original data based on some predefined metric. The experimental results showcase that the proposed approach is useful in generating suitable results based on the critical disease outbreak. It also helps doctors to recheck further verification of virus by the proposed method. The outbreak of Coronavirus has the nature of exponential growth and so it is difficult to control with limited clinical persons for handling a huge number of patients with in a reasonable time. So it is necessary to build an automated model, based on machine learning approach, for corrective measure after the decision of clinical doctors. It could be a promising supplementary confirmation method for frontline clinical doctors. The proposed method has a high prediction rate and works fast for probable accurate identification of the disease. The performance analysis shows that a high rate of accuracy is obtained by the proposed method.

## Introduction

It is now known that the coronavirus disease 2019 pneumonia occurred in the city of Wuhan, China, at the end of 2019. The World Health Organization (WHO) identified and named novel coronavirus as “2019-nCoV” which was later declared as Public Health Emergency of International Concern on January 30, 2020 and further on March 11, 2020 this Covid-19 was characterized as Pandemic [1]. Epidemics of two betacoronaviruses, named as, severe acute respiratory syndrome coronavirus (SARS-COV) and Middle East respiratory syndrome coronavirus (MERS-COV) affected more than 10,000 people in cumulative order in the past two decades[2]. According to the Centres for Disease Control and Prevention (CDC), this novel coronavirus has some similarities with SARS-COV and MERS-COV. These diseases are spread through respiratory droplets from one human being to other. Symptoms as fever, cough, and shortness of breath after a period ranging from 2 to 14 days are observed as the outcomes of the disease [3].

Human to human contacts are considered to be responsible for community spread of this disease in exponential growth. Hence social detachment of people should be followed necessarily to combat COVID-19 from the front line as well as in backend also. Control measures are to be imposed from the government side to implement social detachment of people. Preventive measures may include actions such as locking down the countries, closing and/or minimizing travel connections amongst the countries and within the cities as well, enforcing quarantine and hospitalization of infected individuals, suspending schools, offices, shopping malls, restaurants etc. Curfew imposed in the affected countries is facing potentially huge economical loss. As this disease spreads rapidly, timing is an important factor for controlling the disease as early as possible. Hence, from the initial stage an enormous level of monitoring is required for the authorities in order to handle the epidemic situation. All over the world Scientists are doing 24 hours for finding the vaccine of the disease.

For assisting health planning for Covid-19, Machine Learning framework is proposed in this paper. This will confirm the confirmed cases, negative cases, recovered cases, and death cases considering the cases present in the dataset. The proposed prediction model ensures that it follows the original result regarding this epidemic situation so that enormous economic loss, community spread, amount of social detachment of people may be detected and also accurate decision can be taken accordingly. This method will ensure government authorities to yield preventive measures based on our next work for forecasting the occurrence of this disease in future.

In this paper, data mining concepts are exploited for obtaining prediction of confirmed, negative cases, recovered cases, and death cases where Recurrent Neural Network (RNN) is also employed. The real cases and prediction cases are compared based on some predefined metric. A combined model consisting of Long short-term memory (LSTM) and Gated Recurrent Unit (GRU) is applied to the dataset finally for training and testing purpose. A comparative study is drawn amongst the performance of proposed three models-LSTM-RNN, GRU-RNN, and LSTM-GRU-RNN.

### Related Work

In [4], an AI based approach is proposed as an alternative to epidemiological model for monitoring transmission dynamics for Covid-19. This AI based approach is executed by implementing modified stacked auto-encoder model. This model performs real-time forecasting of the confirmed cases of Covid-19 across China. This model is applied on the dataset collected from January, 11 to February 27, 2020 given by World Health Organization (WHO). Use of latent variables in the auto-encoder and clustering algorithms helps in investigating the transmission procedure by grouping the provinces/cities.

In [5], a methodology called Group of Optimized and Multisource Selection (GROOMS) is suggested which ensembles a collection of five groups of forecasting methods. Classical time-series forecasting methods as well as machine learning methods are implemented where small available dataset are passed from top to down through optimization processes. This will prepare the best winning models for panel section with lowest error. A polynomial neural network and corrective feedback (PNN+cf) is assimilated and implemented in this paper. Experimental results indicate that the combined approach PNN+cf outperforms well over other approaches in terms of Root Mean Squared Error (RMSE) performance measure parameter.

A computation and analysis based on Suspected-Infected-Recovered-Dead (SIRD) model is provided in [3]. Based on the dataset available from January 11 to February 10 2020, it estimates of the main epidemiological parameters, i.e. the basic reproduction number (R_0_) and the infection, recovery and mortality rates, along with their 90% confidence intervals are provided. Computations on SIRD model, this R_0_ parameter value turn out to be 2.5. Experimental results forecast declining mortality rate that in turn help government authorities to impose safeguards.

### Performance Measure Metrics

To identify best candidate model from its peers, it is necessary to put concentration on comparison of measures of the algorithm’s performance. In this paper, following parameters are used for measuring algorithm’s performance-

1. **Accuracy** identifies the overall effectiveness of the algorithm. It is formulated as follows [6]-

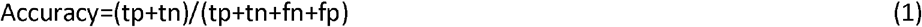

where tp denotes true positive, fp – false positive, fn – false negative, and tn – true negative counts.
2. **Root-mean-square-error (RMSE)** is a standard performance measure used for time series forecasting purpose. It is formulated as follows [7]-

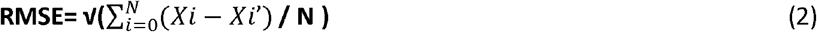

Where Xi is the real value and Xi’ is the predicted value.

### Proposed Models

In this paper, a prediction of confirmed cases, negative cases, released, and deceased cases of Covid-19 corona virus are obtained using a Recurrent Neural Network method. A Recurrent Neural Network (RNN) is kind of neural network architecture that considers both sequential and parallel information processing. Incorporating memory cells to neural network; it is possible to simulate the operations similar to human brain [8]. Following diagram depicts the general structure of a RNN.

Another RNN called Bidirectional RNN (BRNN) accesses future and past context in both directions. There are alternatives from RNN depending on the gating units, such as Long Short Term Memory (LSTM) RNN and Gated Recurrent Unit (GRU) RNN.

Traditional RNN lacks of considering context based prediction, which can be overcome by introducing Long short-term memory (LSTM). LSTM has a good potential to regulate gradient flow and enable better preservation of long-range dependencies [7].

Gated Recurrent Unit (GRU) is quite similar to LSTM, where the gating units of GRU control the flow of information inside the unit, without considering separate memory cells[9]. Like LSTM, GRU lacks of having memory cells in it and it has a lesser number of gates are required and the gates are activated using current input as well as previous output. As compared to LSTM, GRU has better convergence rate due to the reduction of parameters and in some cases GRU outperforms well over LSTM models[10].

For predicting confirmed, negative, released, deceased cases in Covid-19, dataset from kaggle[11] contains cases from 20^th^ January 2020 to 12^th^ March 2020, are used for training and testing three models.

At first, Data are pre-processed by eliminating missing values, irrelevant values. Then data transforming operations are performed so that it can be given as input to the Deep Learning Models. In this paper, three models are implemented and applied on the dataset for verifying the given prediction results with respect to available data set. The prediction results are measured with respect to performance measure metrics such as-accuracy and RMSE. The accuracy of these three models can be improved by choosing proper parameter values. The default parameters may not provide the maximum performance. Hyper-parameter setting is necessary to improve the accuracy level. However, the RMSE value should be optimised as to signify a better model. It is to be noted here that the dataset contains cases for confirmed, negative, released, dead patients. It is tested in clinical laboratory in presence of clinical doctors. This methodology is performed for each of these individual cases separately. The Figure 1 denotes the proposed methodology.

**Figure 1.**
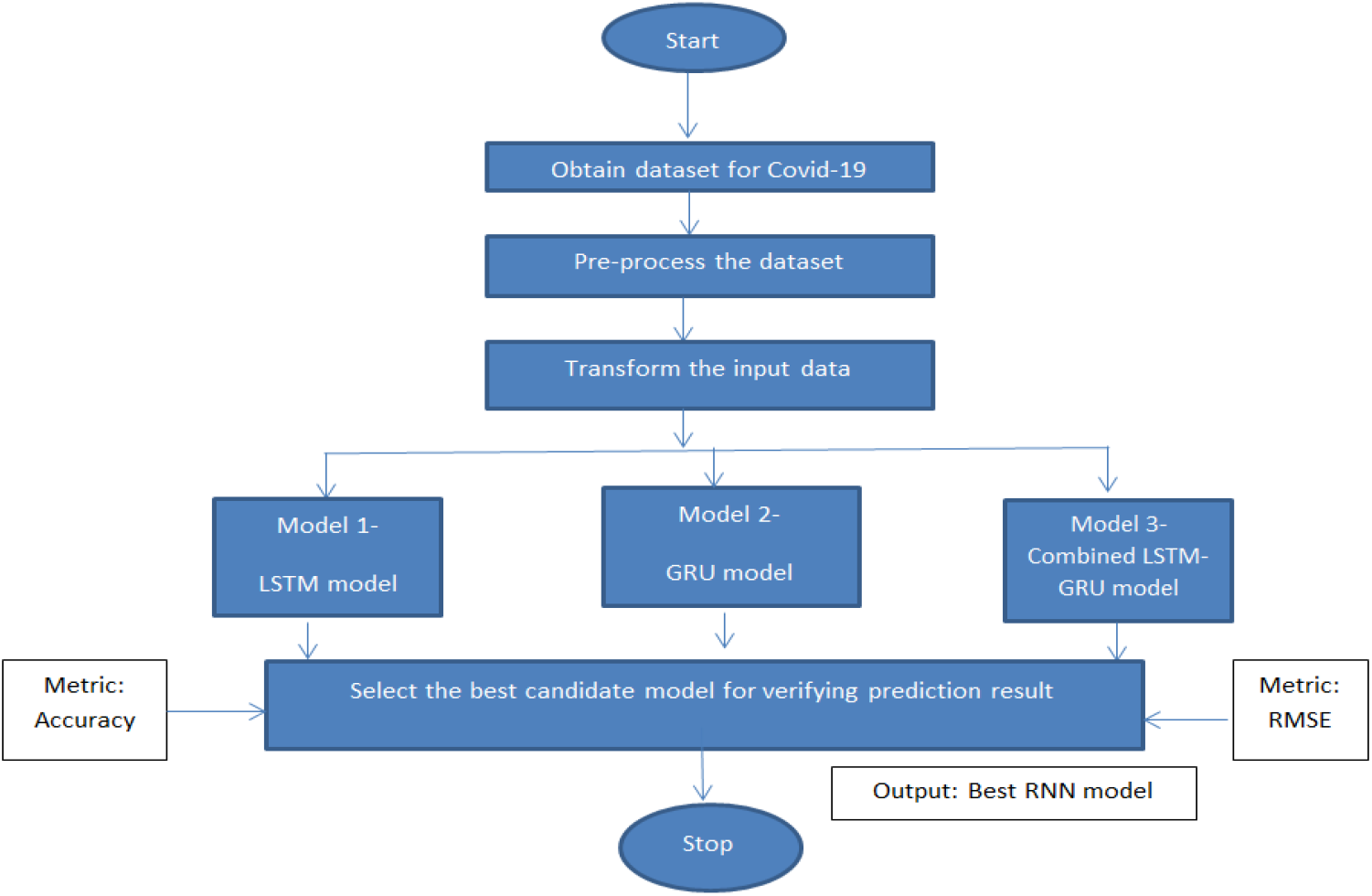
Diagrammatic Approach for the workflow of proposed methodology.

The method consists of three models, which are used for training as well as testing purpose. A detailed explanation with respect to each of these models is provided along with their implementation in the process. The explanation specifies performance of all these models with respect to categories such as, confirmed, negative, deceased, released cases are also presented one after another.

### Model 1

In this model LSTM layers use sequence of 50 nodes. A total 4 layered structure followed by a Dense Layer is used as LSTM model for verifying prediction result. The best hyper-parameters used are a dropout rate of 0.2 and a batch size of 32. The model is shown in Figure 2. The prediction accuracy of the model with respect to each case can be found in Table 1.

**Figure 2.**
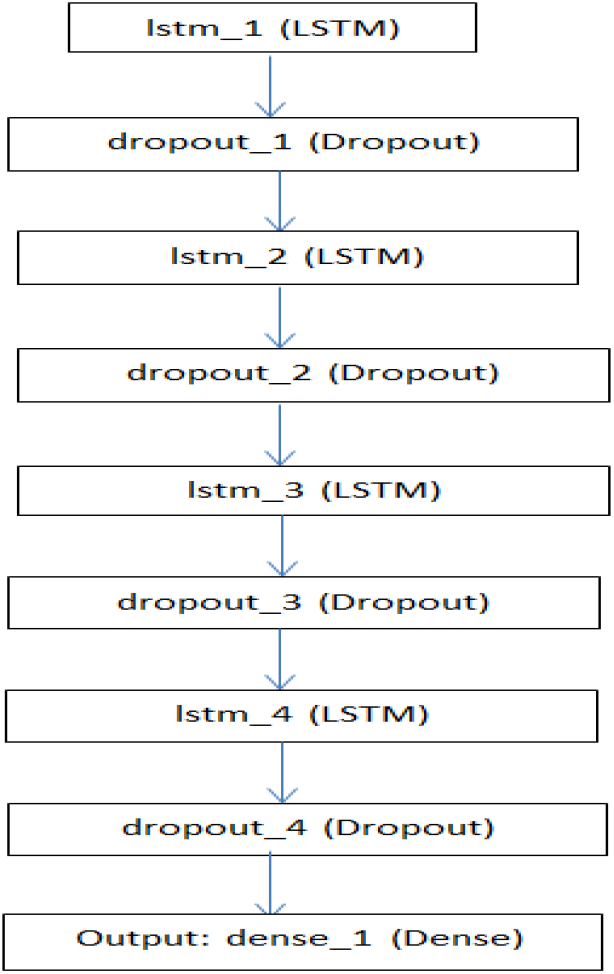
Neural network structure for model 1.

**Table 1.**
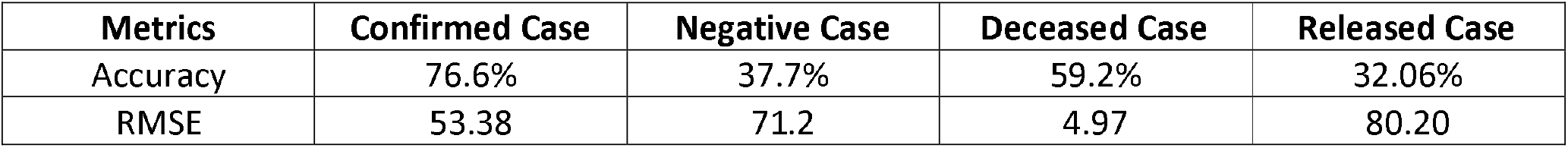
Prediction Accuracy of Model 1.

The accuracy and RMSE depicted in the table may be improved if the model 2 is imposed.

### Model 2

This model has GRU layers and it uses sequence of 50 nodes. A total 4 layered structure followed by a Dense Layer is used as GRU model for prediction result verifying purpose. The best hyper-parameters used were a dropout rate of 0.2 and a batch size of 32. The prediction accuracy of the model can be found in Table 2. A structure of this model is shown in Figure 3.

**Table 2.** Prediction Accuracy of Model 1.

| Metrics | Confirmed Case | Negative Case | Deceased Case | Released Case |
| --- | --- | --- | --- | --- |
| Accuracy | 76.9% | 42.6% | 60.38% | 32.08% |
| RMSE | 30.95 | 70.7 | 5.218 | 80.12 |

**Figure 3.**
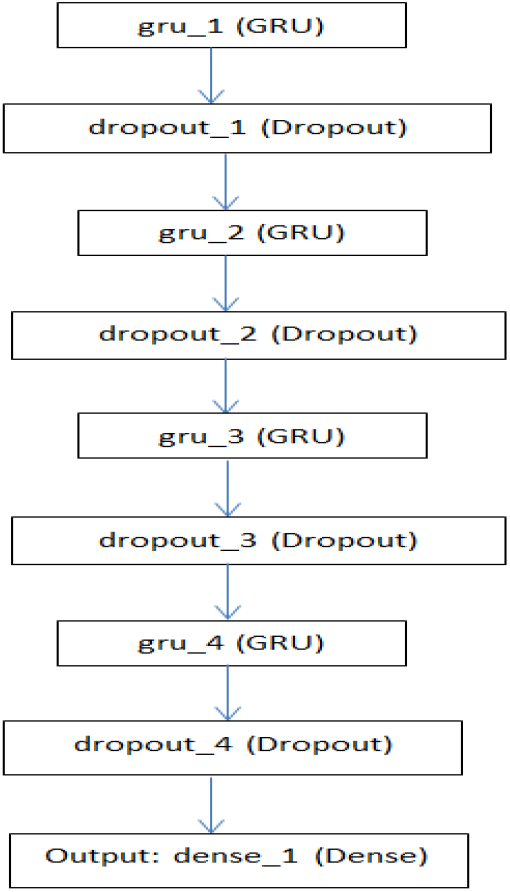
Neural network structure for model 2.

With Respect to the Model 1, Model 2 gives better result in terms of prediction accuracy as well as RMSE. Next, our target is to improve the result obtained using model 3.

### Model 3

A Recurrent Neural Network is employed where LSTM and GRU are assimilated together in order to predict the above mentioned cases. It is shown in Figure 4. For improving the performance of prediction results LSTM and GRU are assimilated and the transformed input is fitted into it. Table 3 signifies that model 3 imposes better impact in verification of the prediction result with respect to its original dataset.

**Figure 4.**
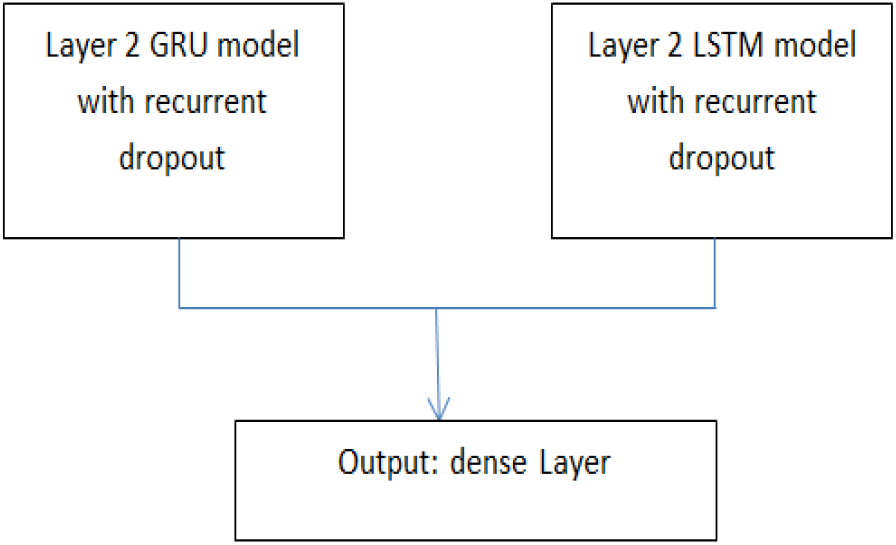
Neural network structure for model 3.

**Table 3.** Prediction Accuracy of Model 3.

| Metrics | Confirmed Case | Negative Case | Deceased Case | Released Case |
| --- | --- | --- | --- | --- |
| Accuracy | 87% | 67.8% | 62% | 40.5% |
| RMSE | 30.15 | 49.4 | 4.16 | 13.72 |

Experimental Results shows that the combined approach LSTM-GRU-RNN provides quite better result over LSTM-RNN, GRU-RNN in terms of Accuracy, RMSE metrics. Figure 5 and Figure 6 depict the overall accuracy as well as RMSE for all these above mentioned model along with the given cases. Higher the accuracy and lower the RMSE, better is the model.

**Figure 5.**
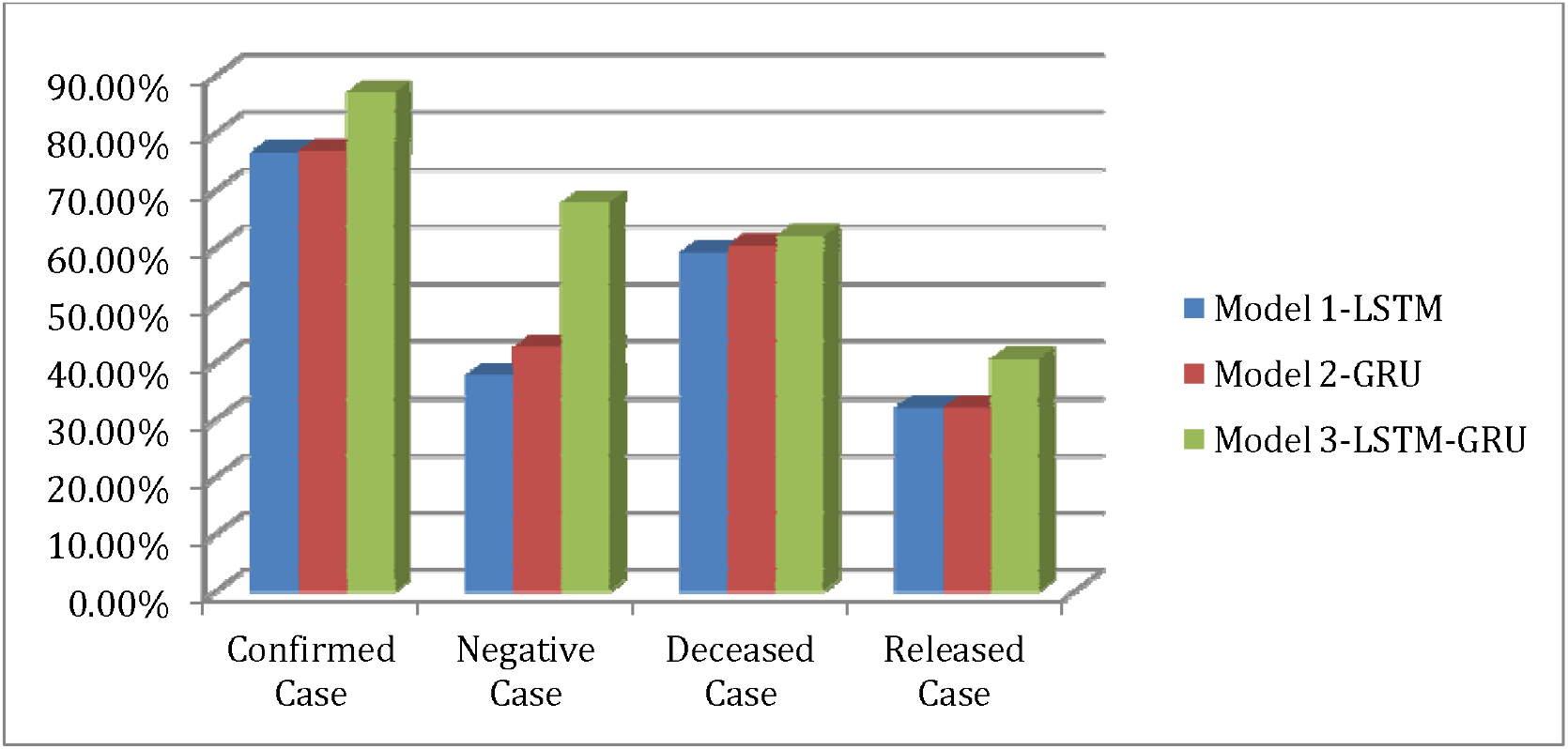
Accuracy Comparison among these three models with along with the cases.

**Figure 6.**
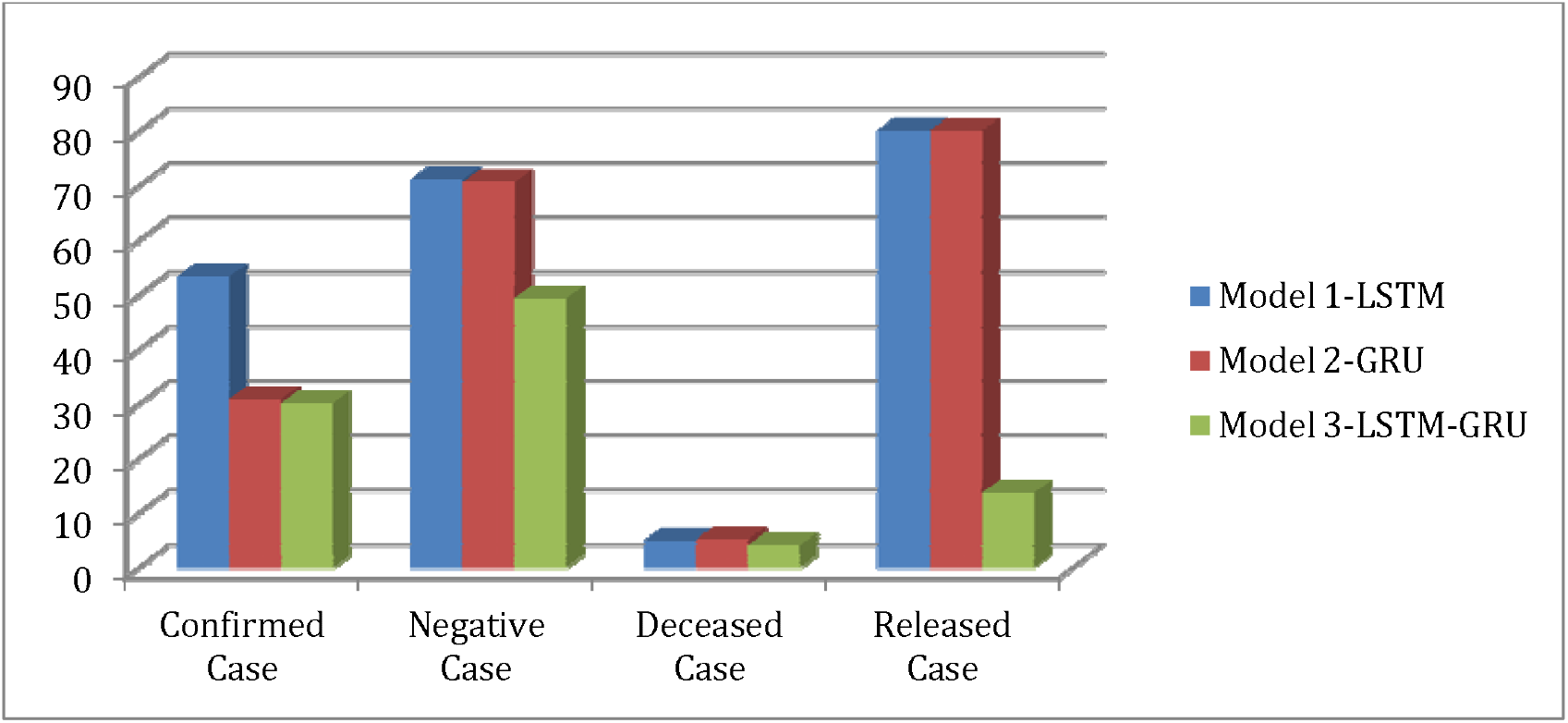
RMSE Comparison among these three models with along with the cases.

### Experimental Results

From the above discussion, the Model 3 provides better result with respect to Model 1 and Model 2. So the model combines LSTM and GRU RNN shows best performance. Figure 7 to Figure 10 depicts the plotting of real result and predicted result for all the cases.

**Figure 7.**
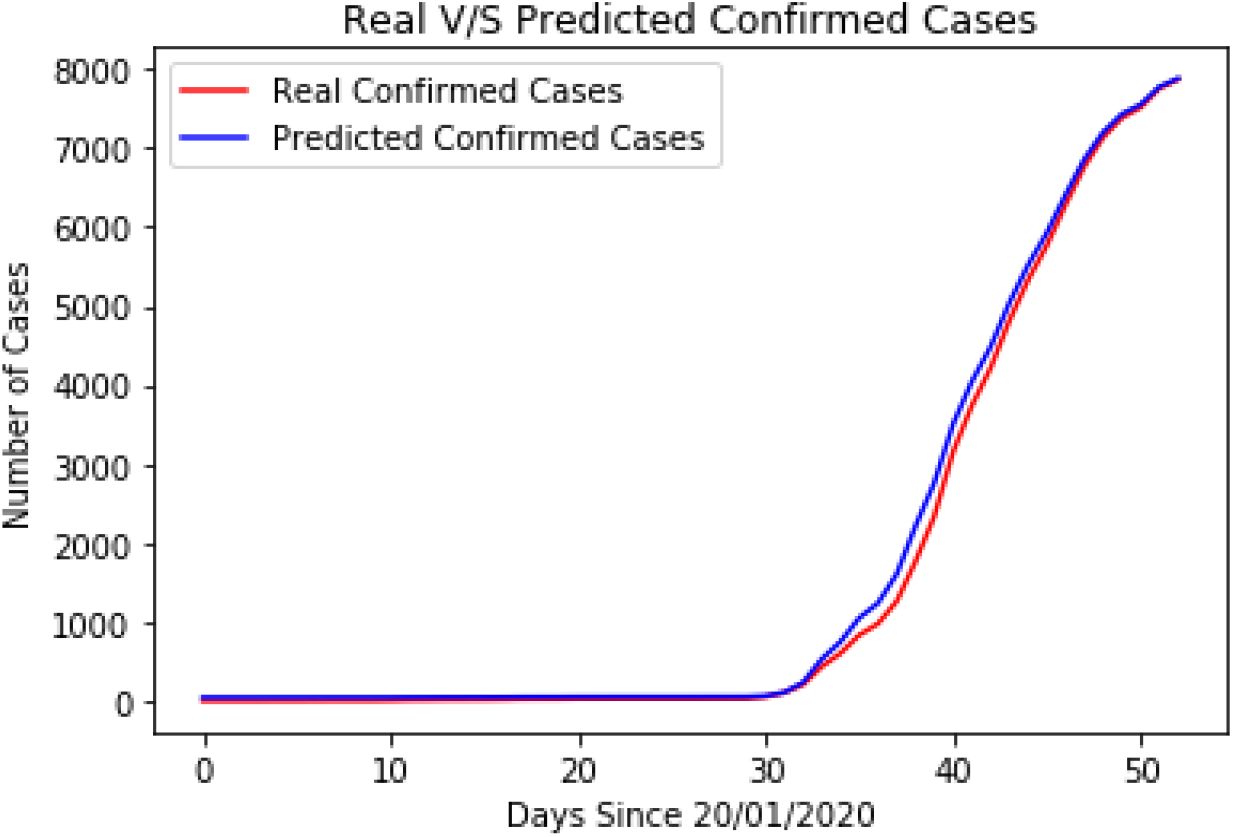
Real V/s Predicted Confirmed Case using LSTM-GRU RNN.

**Figure 8.**
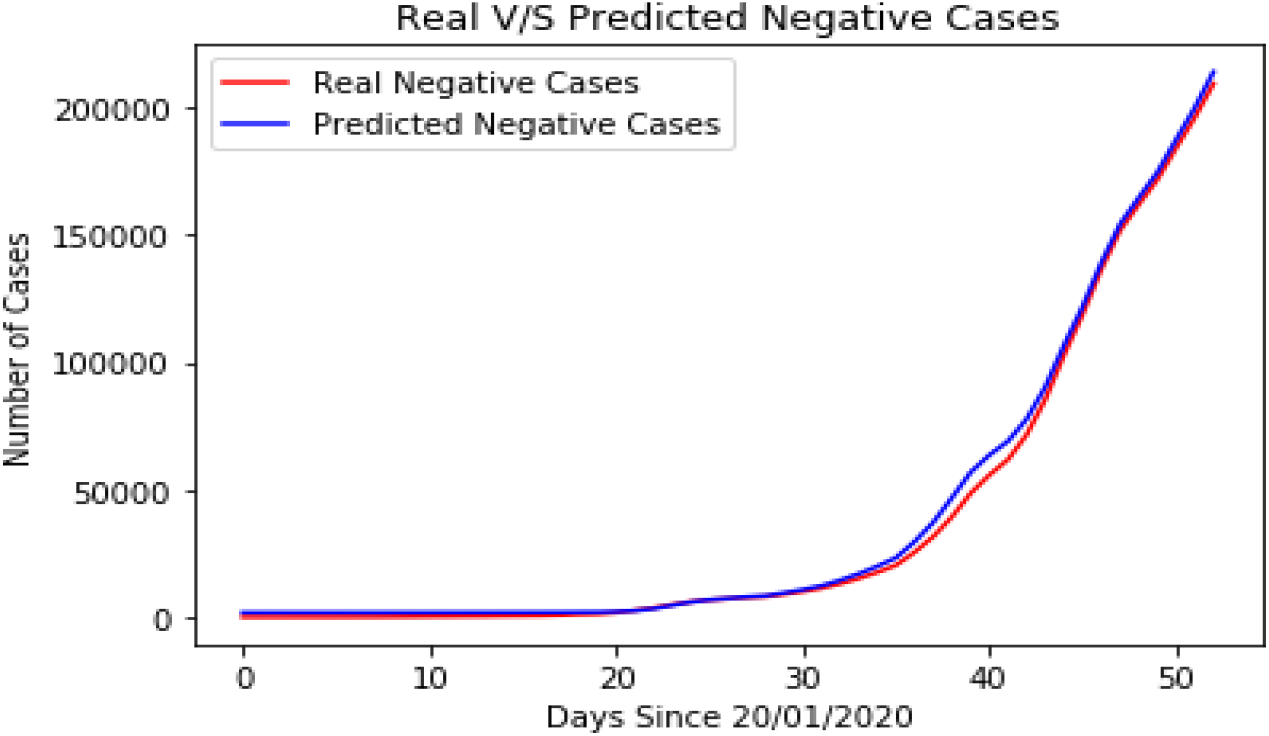
Real case V/s Predicted Negative Case using LSTM-GRU RNN.

**Figure 9.**
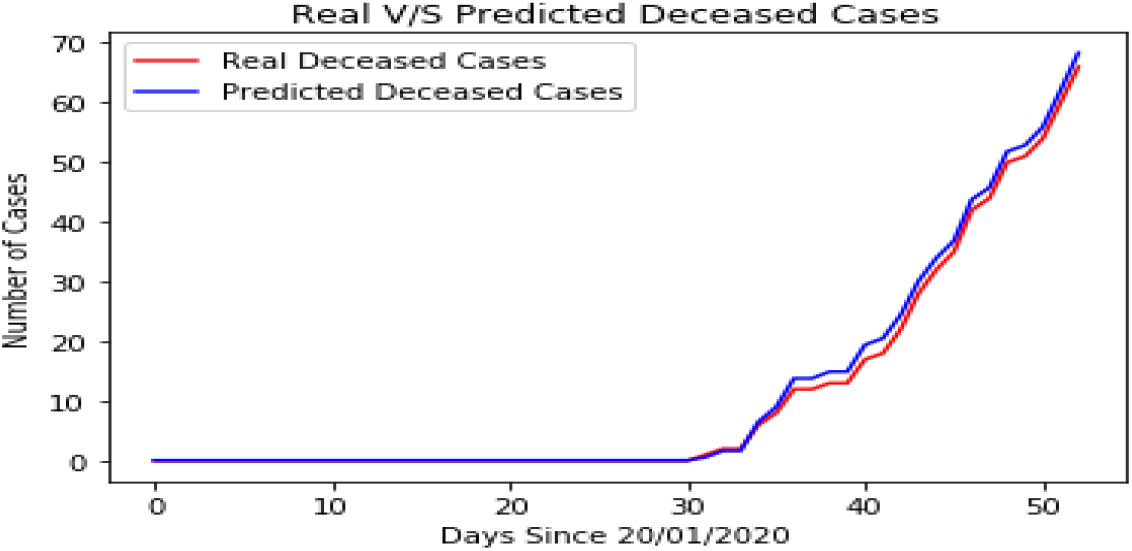
Real V/s Predicted Deceased Case using LSTM-GRU RNN.

**Figure 10.**
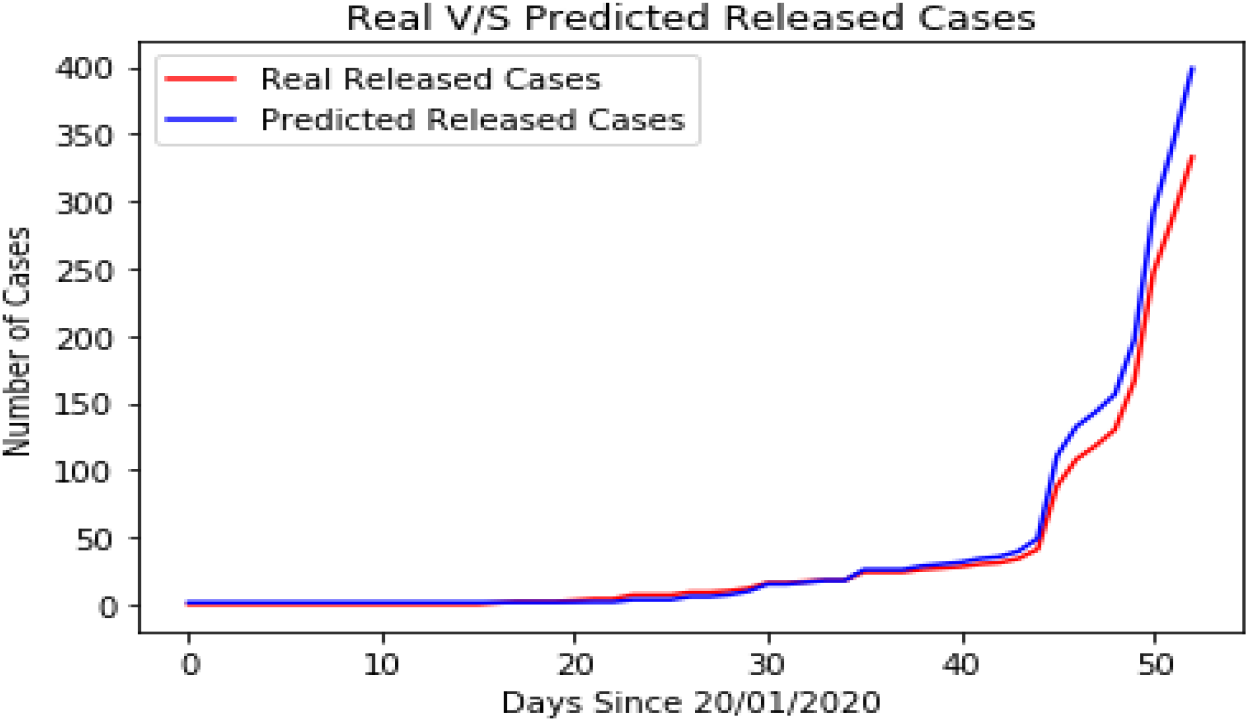
Real V/s Predicted Released Case using LSTM-GRU RNN.

## Conclusions

The combined LSTM-GRU based RNN model provides a comparatively better results in terms of prediction of confirmed, released, negative, death cases on the data. This paper presented a novel method that could recheck occurred cases of COVID-19 automatically. The data driven RNN based model is capable of providing automated tool for confirming, estimating the current position of this pandemic, assessing the severity, and assisting government and health workers to act for good decision making policy. It could be a promising supplementary rechecking method for frontline clinical doctors. It is now essential for improving the accuracy of detection process.

## Data Availability

Datarist (2020, March). Coronavirus-Dataset,Version 1.Retrieved March 17,
2020 from https://www.kaggle.com/kimjihoo/coronavirusdataset-old

https://www.kaggle.com/kimjihoo/coronavirusdataset-old

## Notes

### Competing Interest Statement

The authors have declared no competing interest.

### Funding Statement

The Bhawanipur Education Society College, Kolkata, India

